# Multiregional CT Features Improve Prediction of Immunotherapy Response in Advanced Melanoma

**DOI:** 10.1101/2025.10.26.25338837

**Authors:** Grant Kokenberger, Jatin Singh, Drew Hurd, Tong Yu, Xin Meng, Madison Nguyen, Walter J. Storkus, William McKay, Jason J. Luke, Yana G. Najjar, John M Kirkwood, Hassane M Zarour, Diwakar Davar, Jiantao Pu

**Affiliations:** Department of Radiology, University of Pittsburgh, Pittsburgh, PA 15213, USA; Department of Bioengineering, University of Pittsburgh, Pittsburgh, PA 15213, USA; Division of Hematology-Oncology, Department of Medicine, University of Pittsburgh, Pittsburgh, PA 15213, USA; Department of Ophthalmology, University of Pittsburgh, Pittsburgh, PA 15213, USA

**Keywords:** melanoma, body composition, machine learning, immunotherapy response, CT

## Abstract

**Objective:** Immunotherapy has improved outcomes for advanced-stage melanoma, however, predictive biomarkers remain limited. We evaluated whether computed tomography (CT) features from multiple anatomical regions could predict immunotherapy response.

**Materials and Methods:** This study included 157 advanced cutaneous melanoma patients (mean age: 63.3 years; 65.6% male) treated with PD-1 immune checkpoint inhibitor (ICI) singly or in combination with LAG-3 or CTLA-4 ICIs. The primary outcome was 1-year progression-free survival (PFS ≥12 months). Available artificial intelligence (AI) algorithms were applied to pretreatment CT scans to extract and quantify three-dimensional (3D) body composition and thoracic features across abdominal, chest, pelvic regions, and spinal vertebrae. Feature relationship to PFS was assessed. Machine learning (ML) models were used to predict PFS, utilizing only the most important five features to mitigate overfitting. Prediction performance was evaluated using the area under the receiver operating curve (AUROC) with stratified 10-fold cross-validation.

**Results:** Multiple CT features are significantly associated with immunotherapy response. Body tissues at the L5 spinal vertebrae emerged as key predictors. A random forest classifier (RFC) trained on three CT features (L5 bone volume, L5 subcutaneous adipose tissue volume, pelvis visceral adipose tissue density) and two clinical variables achieved a mean AUROC of 0.83 (95% CI: 0.72–0.94). A logistic regression model using the same features yielded an AUROC of 0.82 (95% CI: 0.74–0.91), significantly outperforming a clinical-only model (AUROC 0.65; 95% CI: 0.56–0.74; p=0.006).

**Conclusion:** Integrating imaging features from multiple anatomical regions can improve the prediction of immunotherapy response in advanced melanoma.

## 1. Introduction

Programmed death-1 (PD-1) is a receptor expressed by activated T cells that ligates PD-L1 (1) and PD-L2 (2) to negatively regulate adaptive immune responses. Cytotoxic T lymphocyte-associated antigen-4 (CTLA-4) is an activation-induced glycoprotein that down-regulates T cell responses by acting as a decoy receptor (3, 4). Immune checkpoint inhibitors (ICIs) targeting these negative regulatory checkpoints have improved response and survival rates in multiple malignancies, including advanced melanoma. In melanoma, the therapeutic efficacy of ICIs is particularly notable, with phase III clinical trials (e.g., CheckMate-067, KEYNOTE-006) reporting objective response rates of 55% with an overall survival rate of 43% at 10 years for the ipilimumab and nivolumab combination (5). Despite these advances, a substantial proportion of patients do not respond to ICIs, highlighting the need to identify predictive markers of treatment response.

Multiple tumor microenvironment (TME) biomarkers have been identified as potential indicators of response to ICIs, including CD8+ TIL-infiltrate, PD-L1 expression, and tumor mutation burden (TMB)(6). However, it is recognized that TME-centric factors alone are insufficient for accurately and reproducibly predicting ICI response. Growing evidence suggests that additional host factors, including HLA class I haplotype, circulating exhausted-phenotype CD8+ T cells, and plasma proteomic profiles, also play significant roles in treatment outcomes. While individual biomarkers offer some predictive value, their effectiveness varies across cancer types and is often limited when used in isolation.

Standard-of-care computed tomography (CT) imaging has a high value potential due to its wide availability, non-invasive nature, and the wealth of information it provides (7–11). CT scans provide detailed images of various tissues and structures, including body composition, which reflects nutritional status and plays a crucial role in immune system health. Variations in adiposity and muscle can alter the body’s metabolism and drug distribution, potentially influencing drug effectiveness and side effects (10). A meta-analysis by Kuang et al. (10) linked low skeletal muscle index (SMI), high subcutaneous adipose tissue (SAT) density, and sarcopenia with both poor progression free survival (PFS) and overall survival (OS). Mengoni et al. (11) identified low SAT abundance and density as significant predictors of PFS (p=0.02) in 100 ICI-treated patients. Despite these insights, limitations remain. Most studies used a single abdominal axial CT slice at the L3 vertebra, which, while computationally efficient, assumes a uniform distribution of body composition throughout the body. Additionally, commonly used indices like fat or skeletal muscle area often lack standardized cutoff values, making the classification of abnormal muscle mass or adiposity inconsistent (12).

Unlike the previous studies, we conducted a detailed analysis of body composition using 3D CT scans to identify biomarkers from pretreatment scans that are significantly associated with immunotherapy response, defined as PFS-12 (radiographic non-progression at the 1-year mark). <u>This study has several unique characteristics.</u> First, we implemented a comprehensive segmentation of body composition-related tissues across all CT images, calculating 3D metrics such as tissue density in Hounsfield Units (HU) and volume (in liters). This approach offers a more granular and precise characterization of body composition compared to conventional methods. Second, we divided the body into four distinct anatomical regions, including abdominal, chest, pelvis, and spine. This regional analysis not only maximizes the utility of heterogeneous CT data, which can be acquired from various body regions, but also enables a more targeted understanding of how specific areas of body composition influence immunotherapy outcomes. Third, several machine learning methods are used to integrate significant image features and clinical variables into a computational model for predicting the response to immunotherapy in advanced melanoma.

## 2. Materials and Methods

### 2.1 Study cohort

We identified a cohort of 158 advanced cutaneous melanoma patients from the University of Pittsburgh Medical Center who received ICI treatment between 2015 and 2024 (**Table 1**). Each patient had at least one pretreatment CT scan. Follow-up CT scans were taken on varying dates after treatment. One outlier patient (BMI > 50) was excluded. All patients were treated in the advanced/metastatic setting and received either anti-PD-1 ICI (57/157), anti-PD-1+anti-CTLA-4 ICIs (71/157), or anti-PD-1+anti-LAG-3 ICIs (29/157). The primary analysis set for multivariate analysis and prediction modeling excluded patients with CT scans that did not encompass the entirety of 3 or more body regions (39/157). This left 118 patients with either complete regional coverage (83/118) or patients with 2 or fewer incompletely scanned body regions (35/118). The doses and schedules of treatment were identical to the initial clinical trial reports. In all three cohorts, patients continued treatment till disease progression or the development of unacceptable immune-related adverse events (irAEs). In responding patients, treatment was truncated at 2 years. Response to therapy was assessed using RECIST v1.1 (13). Progression-free survival (PFS) was defined as the time from treatment initiation to disease progression or death. This study was approved by the University of Pittsburgh Institutional Review Board (IRB #: STUDY 21020214).

**Table 1.** Subject characteristics and PFS (n=157)

| Characteristic | Statistic | PFS > 12 | PFS < 12 | P-value |
| --- | --- | --- | --- | --- |
| **Age (Mean ± SD)** | 63.3 ± 14.7 | 66.3 ± 10.8 | 59.4 ± 17.8 | **0.016** |
| **Male Gender** | 103/157 (65.6%) | 58/87 (66.7%) | 45/70 (64.3%) | 0.886 |
| **Height (in)** | 67.03 ± 3.43 | 66.69 ± 3.39 | 67.47 ± 3.46 | 0.219 |
| **Weight (kg)** | 85.62 ± 17.72 | 86.07 ± 17.56 | 85.06 ± 18.03 | 0.703 |
| **BMI** | 29.47 ± 5.47 | 29.97 ± 5.60 | 28.85 ± 5.29 | 0.437 |
| < 20.0 | 2/157 (1.3%) | 0/87 (0.0%) | 2/70 (2.9%) | 0.999 |
| 20.0 to 24.9 | 37/157 (23.6%) | 18/87 (20.7%) | 19/70 (27.1%) | 0.449 |
| 25.0 to 29.9 | 53/157 (33.8%) | 34/87 (39.1%) | 19/70 (27.1%) | 0.161 |
| ≥ 30.0 | 65/157 (41.4%) | 35/87 (40.2%) | 30/70 (42.9%) | 0.866 |
| **ICI** |  |  |  |  |
| PD1-CTLA4 | 71/157 (45.2%) | 32/87 (36.8%) | 39/70 (55.7%) | **0.027** |
| PD1 | 57/157 (36.3%) | 38/87 (43.7%) | 19/70 (27.1%) | 0.048 |
| PD1-LAG3 | 29/157 (18.5%) | 17/87 (19.5%) | 12/70 (17.1%) | 0.859 |
| **Stage** |  |  |  |  |
| Linear (1-5) | 4.57 ± 1.15 | 4.49 ± 1.10 | 4.66 ± 1.20 | 0.375 |
| III-c/d | 8/157 (5.09%) | 5/87 (5.75%) | 3/70 (4.28%) | 0.195 |
| IV-a | 22/157 (14.0%) | 11/87 (12.6%) | 11/70 (15.7%) | 0.749 |
| IV-b | 29/157 (18.5%) | 22/87 (25.3%) | 7/70 (10.0%) | **0.025** |
| IV-c | 67/157 (42.7%) | 34/87 (39.1%) | 33/70 (47.1%) | 0.394 |
| IV-d | 31/157 (19.7%) | 15/87 (17.2%) | 16/70 (22.9%) | 0.498 |
| **Inflammatory Markers (Mean ± SD)** |  |  |  |  |
| ANC | 4.90 ± 2.40 | 4.54 ± 2.12 | 5.35 ± 2.67 | **0.018** |
| ALC | 1.45 ± 0.62 | 1.48 ± 0.54 | 1.41 ± 0.72 | 0.138 |
| NLR (ANC/ALC) | 4.02 ± 2.90 | 3.44 ± 2.58 | 4.75 ± 3.13 | **0.002** |
| **Genetic Testing** |  |  |  |  |
| NF1 (WT) | 77/105 (73.3%) | 33/58 (56.9%) | 44/47 (93.6%) | **<0.001** |
| NRAS (WT) | 84/110 (76.4%) | 44/62 (71.0%) | 40/48 (83.3%) | 0.198 |
| BRAF (WT) | 63/119 (52.9%) | 35/66 (53.0%) | 28/53 (52.8%) | 0.870 |
| **Adv Rx** |  |  |  |  |
| Pembrolizumab | 24/157 (15.3%) | 18/87 (20.7%) | 6/70 (8.6%) | 0.061 |
| Nivolumab | 16/157 (10.2%) | 12/87 (13.8%) | 4/70 (5.7%) | 0.107 |
| Ipilimumab + Nivolumab | 59/157 (37.6%) | 29/87 (33.3%) | 30/70 (42.9%) | 0.290 |
| Relatlimab + Nivolumab | 27/157 (17.2%) | 17/87 (19.5%) | 10/70 (14.3%) | 0.513 |
| Other | 31/157 (19.7%) | 11/87 (12.6%) | 20/70 (28.6%) | **0.022** |
| SD – standard deviation, WT – wild type, Adv Rx – advanced treatment, ICI – immune checkpoint |  |  |  |  |

### 2.2 Imaging Protocols

The CT scans were acquired after intravenous injection of non-ionic iodinated contrast medium ISOVUE 300 using 64 detector row CT scanners LightSpeed VCT (GE Medical Systems, Milwaukee, WI) and Discovery STE (GE Medical Systems, Milwaukee, WI) with the following parameters: automatic tube current ranging from 150mAs to 600mAs; matrix of 512×512; section thickness of 0.625mm to 3.75mm; 64 x 0.625-mm detector collimation; spiral pitch factor of 1.375. The extent of body coverage in patient scans was heterogeneous, with some patients undergoing full-body imaging, while others received scans limited to chest or abdominal regions.

### 2.3 CT Image Features

The average time from pretreatment CT scans to treatment initiation was 28 days, in line with the performance of pretreatment CT imaging in regulatory trials of immunotherapy. Five body composition tissues, including visceral adipose tissue (VAT), SAT, intermuscular adipose tissue (IMAT), skeletal muscle (SM), and bone, were segmented using a previously developed 3-D convolutional neural network (3-D CNN) (14) (15). To normalize anatomical variations across individuals and to address CT scan heterogeneity, body composition tissues spanning the top and bottom of key anatomical regions (abdomen, chest, pelvis, lumbar vertebrae) were quantified (**Figure 1**). The feature metrics for each tissue include volume (measured in liters) and average density in HU. Feature measurements were standardized by the ratio of tissue volume to region cavity volume. We note that these AI algorithms have been extensively validated across multiple studies involving varying CT protocols and scanners, demonstrating reliable performance, and their detailed descriptions can be found elsewhere (14–19). **Figure S1** details the distribution of cases and their segmented regions. For CT scans with complete coverage of the chest region, lung volume (right and left), thoracic cavity volume, and heart volume were computed using our available AI algorithms (17, 20). Additionally, pulmonary arteries and veins were distinguished, allowing for separate volume calculations for each (16).

**Figure 1.**
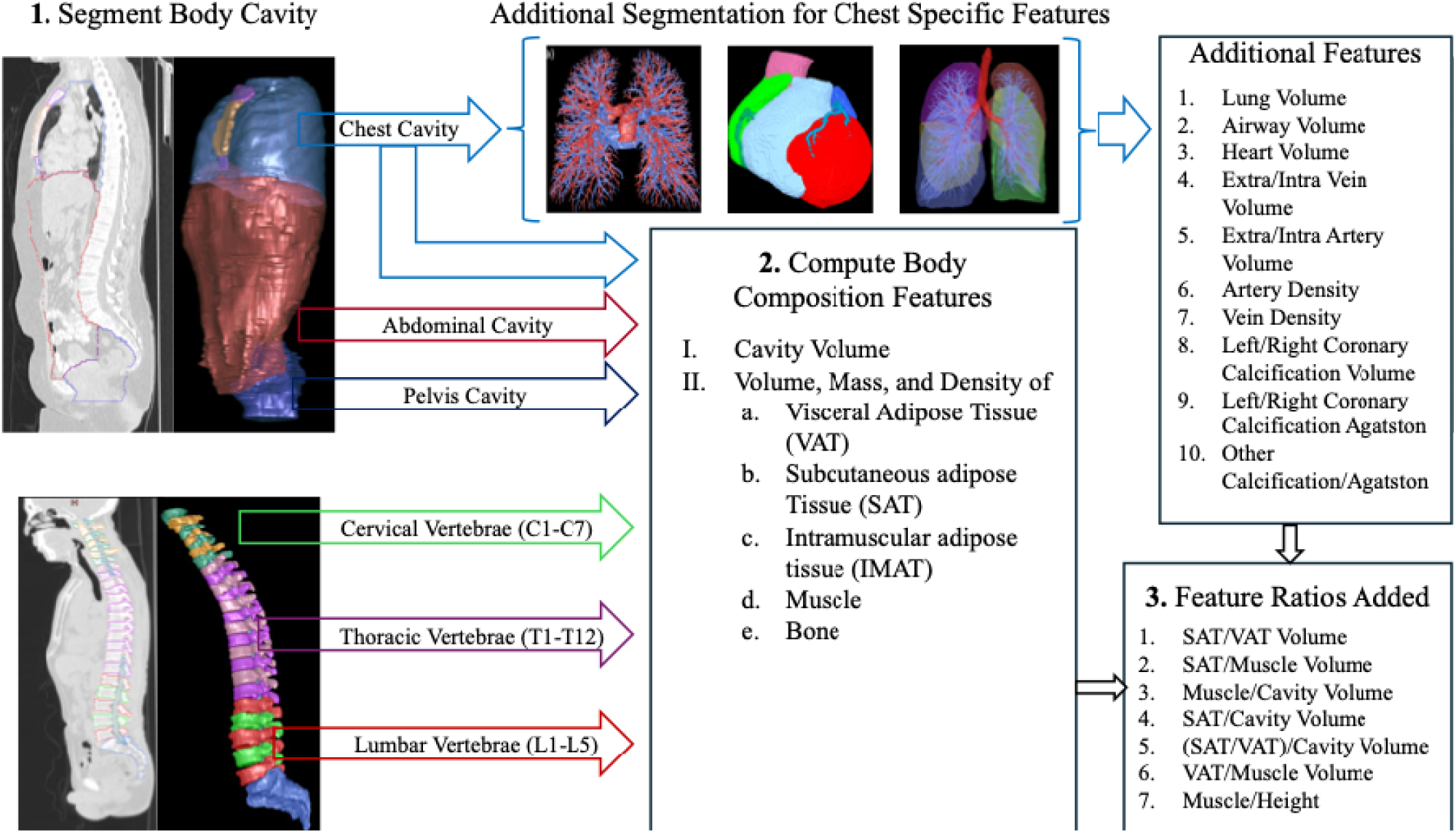
CT-Scan Annotation and Feature Deprivation.

### 2.4 Clinical Features

Pre-treatment complete blood counts (CBCs) were obtained to determine the absolute neutrophil count (ANC) and absolute lymphocyte count (ALC), which were used to calculate the neutrophil-to-lymphocyte ratio (NLR). Follow-up measurements were taken at week 3 to calculate the NLR. Targeted next-generation sequencing (NGS) was conducted using the Oncomine V3 (0.29Mb) assay. DNA was extracted from tumors, and barcoded libraries were prepared for sequencing of hotspots, exons, promoter regions, and introns across 161 genes. Sequencing was performed on an Ion S5 XL System (Thermo Fisher Scientific), and data were analyzed using Torrent Suite Software v5.8.17 and an in-house bioinformatics tool, Variant Explorer (UPMC), to detect point mutations, small insertions/deletions, and copy number alterations.

Variant classification followed the 2017 AMP/ASCO/CAP guidelines(21) using a tier-based system. The assay had a minimum depth of coverage of 300×, with a detection limit of 5% mutant allele frequency (AF). However, only mutations with AF >10% were used for tumor mutational burden (TMB) calculations, excluding variants with AF between 45%-55% due to potential germline origin. The UPMC Oncomine TMB formula accounted for the small panel size using a weighted approach, incorporating variant classification, genomic location, allelic fraction, and minor allele frequency in population databases.

### 2.5 Statistical Analysis

Univariate analysis was conducted for the abdominal, chest, pelvis, and spine subgroups to assess each clinical and pre-treatment CT-derived feature individually. Categorical variables were numerically encoded, and continuous variables were standardized using z-scores. Missing data was imputed using the feature mean. Treatment response determination was defined as a binary variable (PFS-12). The relationship between clinical categorical variables and PFS was tested using Logistic Regression and the chi-squared test. Logistic Regression was also utilized to examine the relationship between linear clinical and CT features and binary PFS. A p-value less than 0.05 was considered statistically significant. Univariate analysis was performed using Python (version 3.13.0).

### 2.6 Prediction Modeling and Performance Evaluation

Four machine learning (ML) models, including logistic regression (LR), support vector machine (SVM), random forest classifier (RFC), and multilayer perceptron (MLP) models, were used and compared for multivariate analysis. Similarly to the univariate analysis, patient CT-derived features were standardized using z-score, and missing data was imputed using the mean. Feature selection was performed using the least absolute shrinkage and selection operator (LASSO) regularization, iteratively increasing the alpha parameter until only the most important 4-5 features remained, as independent testing indicated this feature count optimized performance (**Figure S2**).

A stratified 10-fold cross-validation was used to fit and train each model, ensuring equal distribution of binary classes in each fold. A randomized grid search was conducted on the training set within each fold to fine-tune hyperparameters, including regularization settings, improving model generalization and performance. Model performance was evaluated using the F1 score, the area under the receiver operating characteristic curve (AUROC), and the area under the precision-recall curve (AUPRC). The two-sided Student’s t-test was used to determine statistical significance between the performance scores of clinical, CT, and composite models. Three separate models were trained for each feature subset: one with only clinical and demographic features (Clinical Model), one with only CT-derived features (CT Model), and one with all features (CT/Clinical Model). Feature importance was assessed using SHAP (Shapley Additive Explanations) values in Python. All multivariate analysis was performed using Python version 3.13.0.

## 3. Results

### 3.1 Clinical Features and PFS-12

Advanced age was significantly associated with improved PFS (p=0.016). In line with clinical trial data, the addition of anti-CTLA-4 ICI improved PFS relative to anti-PD-1 ICI monotherapy (p=0.027). However, this effect was not observed with anti-LAG-3 (state p), likely due to the small sample size of this cohort. Expectedly, elevated neutrophil-to-lymphocyte ratio (NLR) was an adverse predictor of PFS (p=0.002).

### 3.2 CT features and PFS-12

Lower intrapulmonary vein volume (mL) from the chest (p=0.047) and lower VAT/SAT ratio in the pelvis (p=0.043) were significantly associated with improved PFS. (**Table 2**). Spinal metrics at the L5 vertebra in the lumbar spine emerged as strong predictors of survival outcomes. Specifically, higher L5 SAT volume (p=0.002), L5 IMAT volume (p=0.007), and L5 muscle volume (p=0.005) were all associated with improved PFS-12. Similarly, among thoracic vertebrae, lower density T5 VAT (p=0.038), greater T7 SAT (p=0.038), T12 SAT volume (p=0.003), and T12 bone volume (p=0.022) were similarly predictive of improved PFS-12. These findings suggest distinct prognostic patterns: greater SAT at thoracic levels and preserved muscle volume at the L5 vertebra are favorable markers, whereas an elevated VAT/SAT ratio may indicate worse outcomes.

**Table 2.** Univariate association of CT features with PFS > 12 (n=157)

| Characteristic | Statistic | OR (95% CI) | PFS > 12 | PFS < 12 | P-value |
| --- | --- | --- | --- | --- | --- |
| Abdominal Region CT Features (n=116) |  |  |  |  |  |
| Volume SAT (L) | 9.04 ± 4.52 | 1.055 (-0.032, 0.139) | 9.5 ± 4.8 | 8.5 ± 4.1 | 0.222 |
| Density Muscle (HU) | 34.11 ± 13.67 | 0.984 (-0.044, 0.012) | 32.8 ± 14.5 | 35.7 ± 12.5 | 0.256 |
| Volume SAT ratio | 0.94 ± 0.46 | 1.576 (-0.381, 1.29) | 1.0 ± 0.5 | 0.9 ± 0.4 | 0.286 |
| Density Bone (HU) | 314.36 ± 48.69 | 0.996 (-0.012, 0.004) | 310.1 ± 50.7 | 319.6 ± 46.0 | 0.298 |
| Chest Region CT Features (n=128) |  |  |  |  |  |
| Intra-Vein Volume (mL) | 82.48 ± 33.37 | 0.989 (-0.022, -0.0) | 77.1 ± 32.4 | 89.0 ± 33.6 | **0.047** |
| SAT/VAT ratio | 5.53 ± 4.12 | 1.068 (-0.033, 0.165) | 6.0 ± 5.0 | 5.0 ± 2.7 | 0.193 |
| Intra-Artery Volume (mL) | 96.75 ± 37.35 | 0.994 (-0.016, 0.004) | 93.1 ± 37.7 | 101.2 ± 36.8 | 0.222 |
| Chest Artery Density (HU) | 82.51 ± 66.86 | 1.003 (-0.003, 0.008) | 87.9 ± 72.3 | 76.0 ± 59.6 | 0.316 |
| Pelvis Region CT Features (n=102) |  |  |  |  |  |
| VAT/SAT ratio | 0.27 ± 0.13 | 0.033 (-6.728, -0.101) | 0.3 ± 0.1 | 0.3 ± 0.2 | **0.043** |
| Density VAT | -77.70 ± 12.18 | 1.023 (-0.011, 0.057) | -76.3 ± 11.9 | -79.5 ± 12.4 | 0.188 |
| Volume SAT ratio | 4.26 ± 1.83 | 1.151 (-0.083, 0.365) | 4.5 ± 1.8 | 4.0 ± 1.9 | 0.217 |
| SAT/VAT ratio | 4.44 ± 2.02 | 1.137 (-0.076, 0.333) | 4.7 ± 1.9 | 4.2 ± 2.1 | 0.218 |
| Lumbar and Thoracic Spine CT Features (n=134) |  |  |  |  |  |
| Volume SAT L5 (L) | 3.82 ± 2.17 | 1.423 (0.136, 0.569) | 4.5 ± 2.2 | 3.1 ± 1.8 | **0.001** |
| Volume Bone L5 (L) | 0.95 ± 0.43 | 5.068 (0.625, 2.621) | 1.1 ± 0.3 | 0.8 ± 0.5 | **0.001** |
| Volume Muscle L5 (L) | 2.78 ± 1.41 | 1.526 (0.125, 0.72) | 3.1 ± 1.0 | 2.4 ± 1.7 | **0.005** |
| Volume IMAT L5 (L) | 0.29 ± 0.19 | 23.709 (0.858, 5.474) | 0.3 ± 0.2 | 0.2 ± 0.2 | **0.007** |
| Volume Bone T12 (L) | 0.25 ± 0.10 | 316.111 (0.829, 10.683) | 0.3 ± 0.1 | 0.2 ± 0.1 | **0.022** |
| Volume SAT T7 (L) | 1.38 ± 0.72 | 1.952 (0.038, 1.3) | 1.5 ± 0.8 | 1.2 ± 0.6 | **0.038** |
| Density VAT T5 (HU) | -69.92 ± 34.28 | 0.983 (-0.034, -0.001) | -77.2 ± 19.1 | -61.6 ± 44.7 | **0.038** |
| Volume Muscle T12 (L) | 0.76 ± 0.30 | 4.534 (0.068, 2.955) | 0.8 ± 0.3 | 0.7 ± 0.3 | **0.04** |
| Volume SAT T6 (L) | 1.25 ± 0.65 | 2.014 (-0.009, 1.409) | 1.4 ± 0.7 | 1.1 ± 0.5 | 0.053 |
| VAT – visceral adipose tissue, SAT – subcutaneous adipose tissue, IMAT – intermuscular adipose tissue, vFMR – VAT to muscle ratio, ratio – tissue volume / region cavity volume |  |  |  |  |  |

### 3.3 ML-Based Prediction Models

Due to variations in CT field-of-view (FOV), some anatomical regions (e.g., abdominal, chest, pelvic, or spine) might not be fully captured. Subjects missing three or more scan regions (N=39) were excluded, while those with one or two missing scan regions were included with imputed data (N=35). In total, 118 subjects, each with CT features from one pre-treatment CT scan, were included in model training and testing.

Features included in the clinical, CT, and CT-Clinical composite models can be found in **Figure 2**, with features ranked from most important to least important.

**Figure 2.**
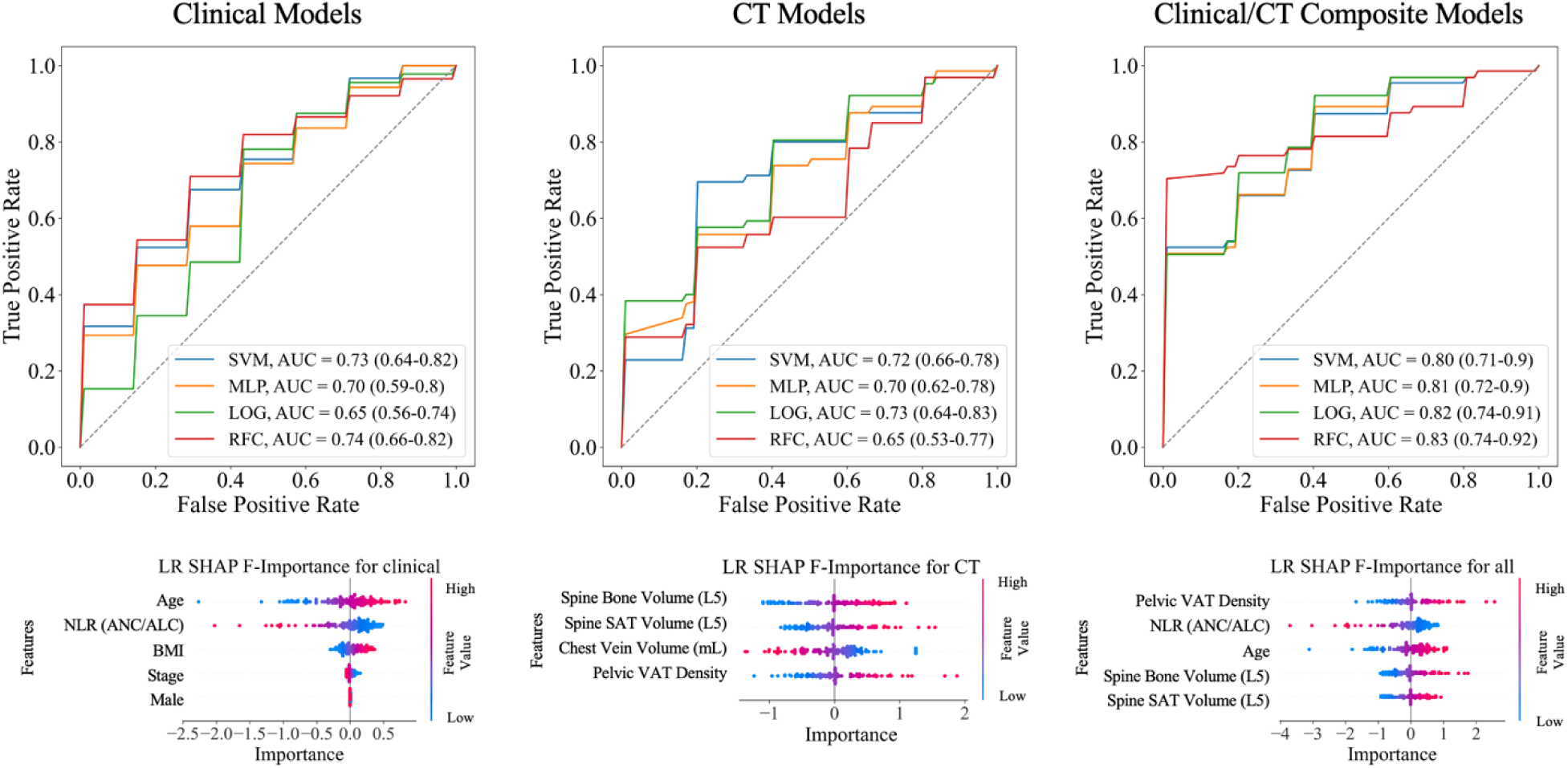
ROC of models using Clinical, CT, and combined Clinical/CT feature subsets, with and SHAP Feature Importance of Logistic Regression Model for each.

Among evaluated models, the random forest classifier (RFC) utilizing the composite CT-Clinical feature subset achieved the highest AUROC 0.83 (95% CI: 0.72–0.94) (**Figure 2**), with statistically significant improvements over the CT-only model (p=0.01) but not the clinical-only model (p=0.099). For the composite models, the RFC model also achieved the highest mean AUPRC (0.90, 95% CI: 0.84–0.95), significantly improving results over CT-only (p=0.004) and clinical-only (p=0.031) models. The logistic regression (LR) model also performed competitively, achieving an AUROC of 0.82 (95% CI: 0.74–0.91) for the composite model, with statistically significant improvements over the clinical model (p=0.006) but not the CT-only model (p=0.121). The MLP and SVM models closely matched these results, with mean AUROC scores of 0.81 (95% CI: 0.72–0.90) and 0.80 (95% CI: 0.71–0.90), respectively, but performance differences were not consistently significant across comparisons (**Table 3**).

**Table 3.**
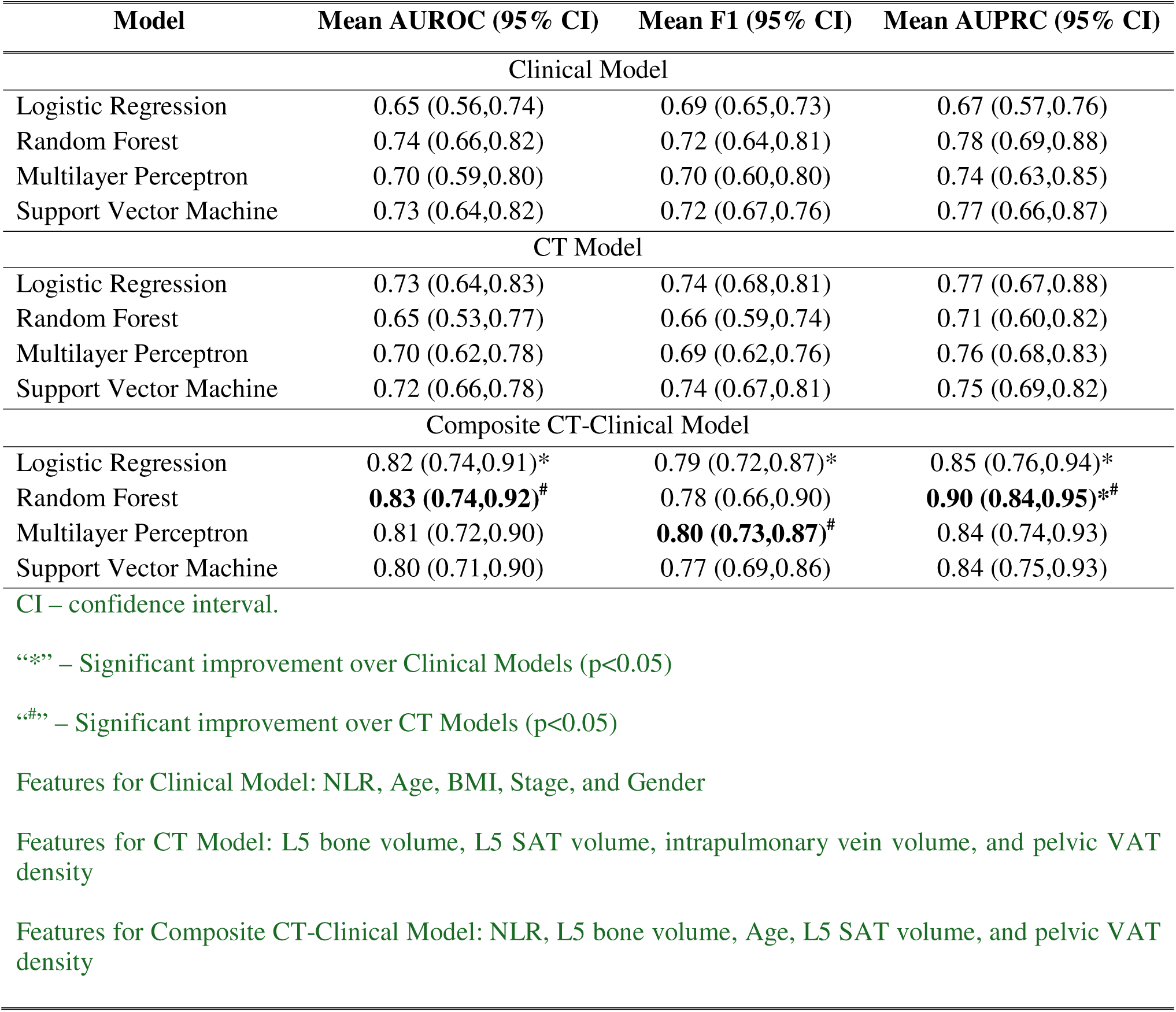
Model performances in predicting PFS > 12 using pre-treatment features (n=118)

| Model | Mean AUROC (95% CI) | Mean F1 (95% CI) | Mean AUPRC (95% CI) |
| --- | --- | --- | --- |
| Clinical Model |  |  |  |
| Logistic Regression | 0.65 (0.56,0.74) | 0.69 (0.65,0.73) | 0.67 (0.57,0.76) |
| Random Forest | 0.74 (0.66,0.82) | 0.72 (0.64,0.81) | 0.78 (0.69,0.88) |
| Multilayer Perceptron | 0.70 (0.59,0.80) | 0.70 (0.60,0.80) | 0.74 (0.63,0.85) |
| Support Vector Machine | 0.73 (0.64,0.82) | 0.72 (0.67,0.76) | 0.77 (0.66,0.87) |
| CT Model |  |  |  |
| Logistic Regression | 0.73 (0.64,0.83) | 0.74 (0.68,0.81) | 0.77 (0.67,0.88) |
| Random Forest | 0.65 (0.53,0.77) | 0.66 (0.59,0.74) | 0.71 (0.60,0.82) |
| Multilayer Perceptron | 0.70 (0.62,0.78) | 0.69 (0.62,0.76) | 0.76 (0.68,0.83) |
| Support Vector Machine | 0.72 (0.66,0.78) | 0.74 (0.67,0.81) | 0.75 (0.69,0.82) |
| Composite CT-Clinical Model |  |  |  |
| Logistic Regression | 0.82 (0.74,0.91)* | 0.79 (0.72,0.87)* | 0.85 (0.76,0.94)* |
| Random Forest | **0.83 (0.74,0.92)<sup>#</sup>** | 0.78 (0.66,0.90) | **0.90 (0.84,0.95)\*<sup>#</sup>** |
| Multilayer Perceptron | 0.81 (0.72,0.90) | **0.80 (0.73,0.87)<sup>#</sup>** | 0.84 (0.74,0.93) |
| Support Vector Machine | 0.80 (0.71,0.90) | 0.77 (0.69,0.86) | 0.84 (0.75,0.93) |

CI – confidence interval.
“\*” – Significant improvement over Clinical Models (p<0.05)
“<sup>#</sup>” – Significant improvement over CT Models (p<0.05)
Features for Clinical Model: NLR, Age, BMI, Stage, and Gender
Features for CT Model: L5 bone volume, L5 SAT volume, intrapulmonary vein volume, and pelvic VAT density
Features for Composite CT-Clinical Model: NLR, L5 bone volume, Age, L5 SAT volume, and pelvic VAT density

### 3.4 Feature Importance

SHAP analysis was used to evaluate the importance of each feature subset (clinical, CT, composite). The top three features from the composite LR model were pelvis VAT density, NLR, and Age (**Figure 2**). The SHAP summary plot provides an interpretable visualization of feature importance and its impact on the model’s predictions. Features are ranked vertically by their overall contribution to the model, with the most influential variables positioned at the top. The x-axis represents SHAP values, which quantify the magnitude and direction of each feature’s effect on the model’s predicted outcome. Additionally, color gradients indicate feature values, where higher values are typically shown in red and lower values in blue.

## 4. Discussion

Identifying patients likely to respond to ICI remains a major challenge in improving survival for advanced melanoma. We investigated the relationship between CT-derived features and ICI-based treatment response in advanced-stage melanoma patients. By analyzing a broad range of body tissues in four distinct body regions, we captured a holistic view of patient health. Additionally, we included CT-based measurement of the lung, heart, and vascular structures to enhance prognostic and predictive accuracy. To address variations in CT protocols and scan fields of view, we normalized body composition metrics by dividing tissue volumes by the corresponding region cavity volumes. Our findings suggest that an optimal combination of body composition measurements from multiple regions can help identify patients who are more likely to benefit from immunotherapy, offering a potential index to avoid unnecessary treatment in non-responders. Unlike other biomarkers, CT-derived body composition features are readily quantifiable from routinely acquired, non-invasive CT scans in cancer care without incurring additional tests and radiation exposure.

A detailed analysis of various body tissues based on CT scans offers greater detail than BMI, which often yields inconclusive or conflicting results, as shown by available studies (22, 23). Prior studies have examined body composition features (10, 11, 24–28) or clinical features(29–33) in relation to immunotherapy outcomes, with some investigations using machine learning prediction methods(8, 9, 28, 34, 35). However, most of these studies relied on metrics derived from single cross-section slices at specific locations (e.g., L3/L4), limiting the accuracy and robustness of the findings (36). Additionally, commonly used indices like fat or skeletal muscle area often lack standardized cutoff values, making the classification of abnormal muscle mass or adiposity inconsistent (12). To overcome these limitations, we quantified 3D body tissues across all image slices within segmented anatomical regions (**Figure 1**), enabling a more accurate and comprehensive body composition assessment. A regional analysis provides a more nuanced understanding of localized tissue characteristics and their clinical relevance. Additionally, relying on single-slice or whole-body measurements can overlook regional tissue characteristics that are more strongly associated with treatment response. While whole-body tissue biomarkers have yet to be firmly established, region-specific features hold significant potential as predictive biomarkers. By focusing on specific anatomical landmarks, regional body composition analysis may reveal distinct patterns of tissue distribution and offer additional biomarkers that may influence treatment response.

A notable practical challenge was the heterogeneity of CT scan coverage, which is often inconsistent across patients. To address this, we quantified body tissues across all image slices within each segmented body region to maximize the use of the available scans and normalize their metrics. Our findings showed that increased SAT in multiple body regions was consistently associated with prolonged PFS, aligning with previous studies linking higher fat levels to better immunotherapy outcomes (24, 26, 27). SAT may exert protective effects by acting as an energy reserve and physical barrier, while also producing pro-inflammatory cytokines and chemokines that enhance immune cell recruitment and activation.

In the pelvic region, a higher VAT/SAT ratio showed a marginally negative association with PFS. This contrasts with the findings by Esposito et al.(37), who reported a higher VAT/SAT ratio associated with improved OS in ICI-treated patients, indicating the need for further investigation. Greater muscle and IMAT volume were also linked to better PFS-12, consistent with earlier studies examining the role of skeletal muscle in predicting patient responses and survival when treated with ICIs (26–28). Lower VAT density was also associated with improved survival, supporting previous findings that higher-density adipose tissues are linked to higher mortality risk, independent of BMI (38). Additionally, higher bone volume was positively associated with PFS-12, potentially reflecting better skeletal health, as bone metastasis is often accompanied by bone loss and poor ICI response volume. Finally, increased intrapulmonary vein volume was associated with decreased survival, suggesting that structural changes in pulmonary circulation may impact overall health and cancer outcomes by affecting blood flow, nutrient delivery, and tumor progression.

While most prior studies have predominantly utilized the L3 or L4 vertebrae to derive body composition metrics (24, 27), our findings based on comprehensive analyses of body composition at all vertebrae suggest that 3D body tissues quantified from all image slices located at L5 emerged as the strongest predictors.

Significant associations were also observed between clinical variables, genetic testing results, and blood tests. Older age was associated with longer PFS, consistent with previous studies that older melanoma patients often respond better to immunotherapy (29). Elevated NLR and ANC were strongly associated with shorter PFS, in line with prior studies (28, 30, 32). Additionally, NF1-mutated melanoma was correlated with improved PFS, supporting findings by Kroll et al.(31), who reported that patients with NF1-mutated advanced melanoma treated with ICIs experienced longer PFS.

Overall, the ML models demonstrated strong predictive performance for PFS using pre-treatment features, with balanced sensitivity to both positive and negative classes. The top-performing clinical-only model achieved an AUROC of 0.74 (95% CI: 0.66–0.82), while the top-performing CT-only model followed closely with an AUROC of 0.73 (95% CI: 0.64–0.83). Composite models outperformed both, with the RFC model achieving an AUROC of 0.83 (95% CI: 0.74– 0.92). The limited variation among the composite ML models likely reflects the relatively small dataset size. Robust feature elimination combined with consistent performance across stratified cross-validation, along with a small number of features in the models, indicates strong resistance to overfitting. Notably, 3 out of the 5 features in the composite model were derived from CT-derived body composition metrics. Genetic testing results were excluded due to their limited predictive value compared to CT-derived features and NLR. While clinical features have traditionally been used in ML models for melanoma, our findings show that the inclusion of body composition features significantly improves performance over clinical models alone (p<0.05).

This study has several limitations. First, the sample size is relatively small, which may limit the generalizability of the findings and increase the risk of overfitting. To address this, we implemented a 10-fold cross-validation strategy and limited the number of features in the models to enhance the reliability and robustness of the results. Second, the limited racial diversity in the dataset may reduce the external validity of our results, as melanoma outcomes can vary across different demographic groups. Additionally, the absence of an external validation dataset is another limitation. The results presented in this study were derived solely from an internal cohort and included multiple treatments, including single and combination ICI therapy. Furthermore, we did not consider tumor-specific characteristics such as size, shape, border, asymmetry, or evolution because our primary emphasis is on the potential of body composition as a biomarker for immunotherapy response in advanced melanoma.

## 5. Conclusion

This study explores the potential of multiregional pre-treatment CT-derived body composition biomarkers to predict PFS in advanced melanoma patients treated with ICIs. We analyzed a cohort of 157 patients, focusing on a comprehensive set of body tissues (e.g., VAT, SAT, IMAT, skeletal muscle, and bone) and their 3D features in different anatomical landmarks (chest, abdomen, pelvis, and spine). Both statistical and machine learning methods were used to assess the relationship between these CT-derived metrics and ICI response. To our knowledge, this is the only study that performed a comprehensive study of the body composition across multiple anatomical regions and showed that predicting the immunotherapy in advanced melanoma could be significantly improved by their optimal combination.

## Data Availability

All data produced in the present study are available upon reasonable request to the authors

## Abbreviations and Acronyms

AUROC: area under the receiver operating characteristic curve
AUPRC: area under the precision-recall curve
M: machine learning
VAT: visceral adipose tissue
SAT: subcutaneous adipose tissue
IMAT: intramuscular adipose tissue
SVM: support vector machine
MLP: multilayer perceptron
RFC: random forest classifier
CI: confidence interval
BMI: body mass index
ICIs: immune checkpoint inhibitors
OS: overall survival
PFS: progression-free survival
RFS: recurrence-free survival
LR: logistic regression
ANC: absolute neutrophil count
ALC: absolute lymphocyte count

## Notes

**Funding:** This work was supported in part by Developmental Research funding through the University of Pittsburgh Melanoma and Skin Cancer SPORE NIH P50 CA254865 and NIH R01CA237277. Sponsors had no involvement in this study.

**Conflicts of Interest:** None declared

### Competing Interest Statement

The authors have declared no competing interest.

### Funding Statement

This work was supported in part by Developmental Research funding through the University of Pittsburgh Melanoma and Skin Cancer SPORE NIH P50 CA254865 and NIH R01CA237277. Sponsors had no involvement in this study.

### Author Declarations

This study was approved by the University of Pittsburgh Institutional Review Board (IRB #: STUDY 21020214).

